# Detection of an imported case of severe Crimean-Congo hemorrhagic fever virus in a patient with comorbidities, Dakar, Senegal 2023

**DOI:** 10.1101/2024.03.04.24303091

**Authors:** Samba Niang Sagne, Ousseynou Sene, Idrissa Dieng, Mamadou Korka Diallo, Amadou Moustapha Ndoye, Yoro Sall, Boly Diop, Oumar Faye, Abdourahmane Sow, Boubacar Diallo, Cheikh Loucoubar, Gamou Fall, Mamadou Aliou Barry

## Abstract

In July 2023, a diabetic from Mauritania was diagnosed with a severe case of Crimean-Congo hemorrhagic fever at a Dakar region hospital, Senegal. The phylogenetic analysis revealed the new strain as a CCHFV reassortant between Genotype I and III, closely linked to strains from Spain, Mauritania, Senegal and South Africa. Genetic variability of CCHF in West Africa underscores the urgent need for enhanced surveillance in West Africa.

## Introduction

The first clinical report of Crimean-Congo hemorrhagic fever (CCHF) in humans was in the Crimea in 1944 and became known as ‘Crimean hemorrhagic fever’; nonetheless, the virus was first isolated in Congo in 1956 and was named ‘Congo virus’; the two names converged as CCHF in 1969 [1]. CCHF is an often fatal viral infection described in about 30 countries, in Africa, Asia and Europe where the primary vectors *Hyalomma* ticks have a wide distribution [2]. Crimean-Congo hemorrhagic fever virus (CCHFV) infection is correlated with several risk factors, including the bites of ticks, the contact with a patient with CCHF during the acute phase of the infection as well aswith blood or tissues from viremic livestock [2]. Nosocomial transmission is possible via infected fluids or accidents involving virus-contaminated equipments [3]. The initial symptoms of CCHF are aspecific and gradually progress to a hemorrhagic phase, which can be fatal with a fatality rate ranging from 10 to 50% [4]. CCHF is characterized by a sudden onset of high fever, chills, severe headache, dizziness, back and abdominal pains. Additional symptoms can include nausea, vomiting, diarrhea, neuropsychiatric, and cardiovascular changes. In severe cases, hemorrhagic manifestations, ranging from petechiae to large areas of ecchymosis can occur[5].

CCHFV belongs to the genus *Orthonairovirus* of the *Nairoviridae* family [1]. CCHFV has a negative tripartite RNA genome with three segments namely S (small), M (medium), L(large) [1]. The current classification of CCHFV includes five genotypes (I-III in Africa, IV in Asia) and V in Europe). Genotype ? no longer exists based on a new classification where the CCHF strains that belonged to genotype IV have been reclassified to the Aigai viral species [7]. In West Africa, the first human cases of hemorrhagic fever linked to CCHF were described in Mauritania in 1983 [7]. Since them numerous outbreak were described in Mauritania while sporadically cases were identified in Senegal [7–9]. Here, we report the longitudinal clinical and biological observations of a CCHF case imported from Mauritania to Senegal in 2023 as well as the genomic characterization of the detected virus.

### The study

In July 2023, a man aged between 50 - 60 years, with type 2 diabetes, was admitted to intensive care in a Dakar hospital for a febrile syndrome, an algic syndrome associated with vomiting and epigastralgia. the onset of symptoms would date back to a week before his admission. The examination revealed a patient with clear consciousness but asthenic with diffuse skin lesions in the form of petechiae and purpura, clinical anemia, tachycardia, all in a context of severe hyperglycemia at 4,86 g/dl with glycosuria and no ketonuria. Biological findings included: a negative malaria RDT, thick drop, C-reactive protein at 28mg/l bicytopenia with hemoglobin at 9,7g/dl, severe thrombocytopenia at 2000/mm3, impaired renal function with creatinemia at 46 mg/l, hyponatremia at 131mEq/l, hypochloremia at 91mEq/l, hepatic cytolysis with SGOT transaminase at 2020 UI/l, SGPT transaminase at 762 UI/l, biological cholestasis, increased direct bilirubin at 9,7 mg/l. The patient received as first treatments broad-spectrum antibiotics, insulin therapy, antipyretics and platelet concentrate transfusions. On the 1st day of hospitalization, externalized hemorrhagic signs were observed in the form of gingivorrhagia and epixtasis, and the patient was placed in isolation. A suspicion of viral hemorrhagic fever (VHF) was raised, and the sample was taken and sent to the Institut Pasteur de Dakar (IPD), with the support of the sentinel syndromic surveillance of Senegal (4S network) in the Prevention Department of the Ministry of Health. At the IPD, viral RNA was extracted from clinical samples using the QIAamp viral RNA mini kit according to the manufacturer’s instructions (Qiagen, Hilden, Germany) and screened for several arboviruses and viral hemorrhagic fevers (VHF) including CCHF by real time RT-PCR [10]. The sample was positive for CCHFV. Unfortunately, the patient died in hemorrhagic shock. Sequencing was performed according to the Illumina RNA prep with enrichment (L) kit reference guide (1000000124435 v01) on an Illumina MiSeq 150 cycle system. The sequencing reads were analyzed using EDGE bioinformatics pipeline (https://www.edgebioinformatics.org/ accessed on August 08, 2023), which generated a single Fasta file for each sample. Nearly complete genomes obtained during this work were submitted to a public nucleotide BLAST database to identify their homologous sequences. They were then combined with a representative subset of CCHFV sequences available in Genbank. Sequences were aligned using the MAFFT program and ML phylogenetic tree build with IQ-TREE with1000 bootstrap replicates for robustness[11,12].

Our study reports a CCHF case imported from Mauritania, detected in July 2023 in a patient with comorbidities (diabetic). Unfortunately, the patient, who had complication, succumbed. Albeit this case exhibits clinical and biological similarities with the initial documented CCHF associated hemorrhagic syndrome in West Africa, identified in Mauritania, as well as with CCHF case involving two tourist in Dakar in 2004 [8,9].

BLASTn analyses revealed that our strain shares a very high nucleotide sequence similarity (more than 98%) with strains isolated from Spain in 2014 and 2016 (ASV45882, ATG31912), Senegal in 1975 (WAD86875), Mauritania in 1984 (ABB30041) and Sudan in 2009 (AEI70581). Phylogenetic analysis revealed that the new CCHF strain is a probable reassortant between Genotype I (L and M segment) and III (S segment) and clutered with the recent known reassortant strain from Koumpentoum, Northern Senegal in 2022 (Koum_SEN_2022) along the three segment[13]. Furthermore, these strains appears to have multiples origins as they form clusters with strain from Spain in 2014 and 2016, Mauritania in 1984, Senegal in 2019 and 2022 (ArD374334, Boki_CCHF_2019) and South-Africa in 1985 (AAZ38665, ABB30048) across the L, M and S segments (Figure 1). Reassortment in bunyaviruses can alter their pathogenicity, exemplified by Ngari virus, a reassortant orthobunyavirus with L and S segment from Bunyamwera virus and the M segment from Batai virus. Although Bunyamwera and Batai viruses typically cause mild disease, Ngari has been associated with hemorrhagic fever cases in East Africa[14,15]. Therefore, it is crucial to investigate the potential impact of this reassortment on the biological properties of the new strains and ascertain its potential involvement in the observed severe clinical manifestations. The patient had travelled from Mauritania, an epidemic country for CCHFV, which further supports the idea of transboundary transmission contributing to CCHF strain exchange between Senegal and Mauritania.

**Figure 1:**
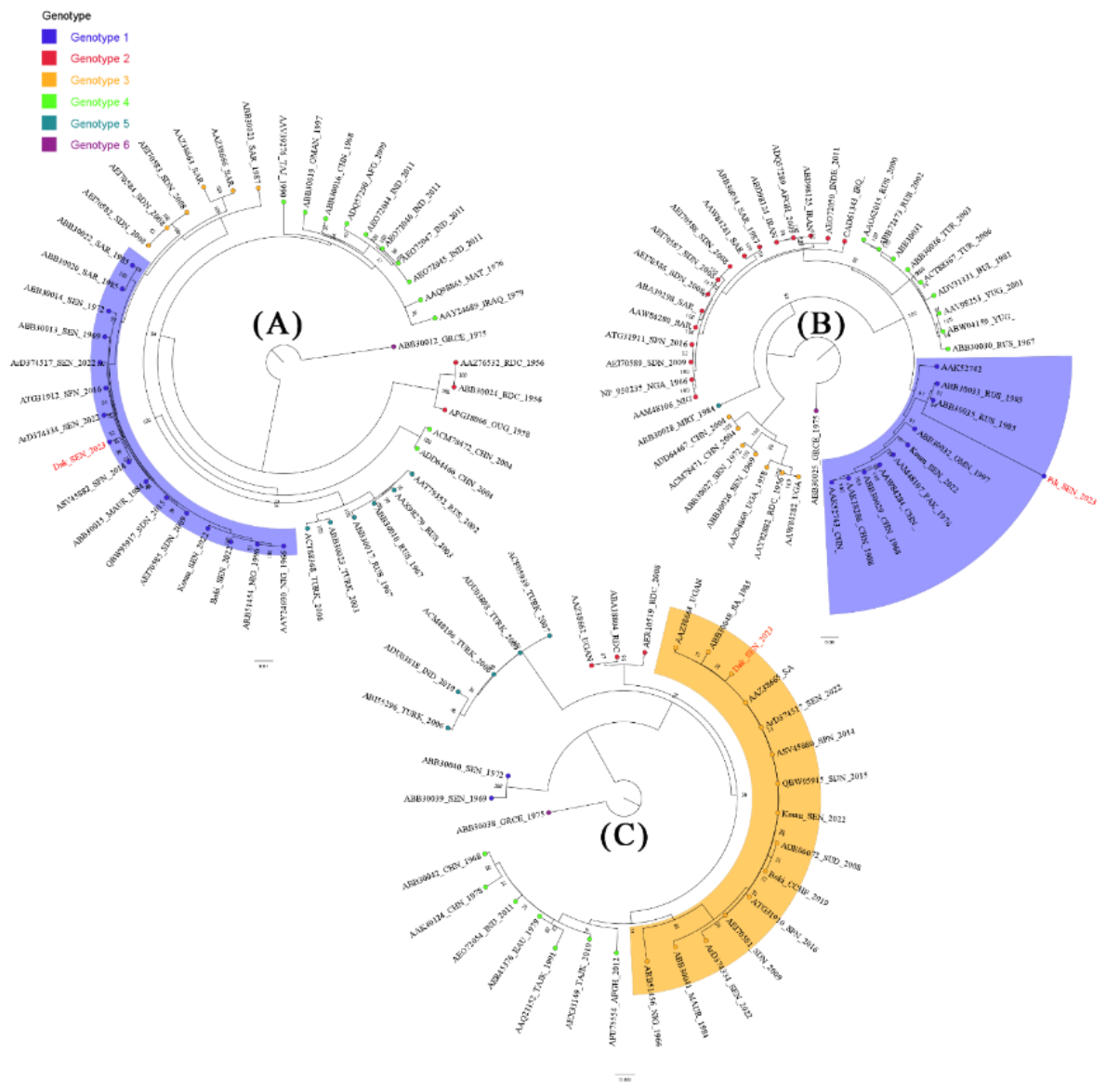
Maximum likelihood trees of CCHFV strains based L, M and S segment (Panel A, B and C). Genotype I is highlight by blue colour and III in orange. The newly characterized CCHF isolates are color-coded in red.

Our study highlights Mauritania’s status as a high-risk area for CCHF and confirm the hypothesis of strains exchange between different countries including Senegal and Mauritania. Additionally, it sheds light on the genetic diversity among West African CCHF strains and raises question about the potential introduction of CCHF strains from other countries including Spain and South Africa and underscoring the need for enhanced surveillance to prevent future outbreaks. It also emphasizes the importance of considering comorbidity factors, such as diabetes, in the management of CCHF cases. The lack of detection of the CCHF case in Mauritania might be due to the unspecific symptoms during the acute stage of the disease or to the absence of an efficient surveillance system. Since October 2023, surveillance has been strengthened in Mauritania by implementing the Senegalese syndromic sentinel surveillance network, that allow early detection of VHF cases in Senegal, through the Africa CDC RISLNET Project.

## Data Availability

All data produced in the present study are available upon reasonable request to the authors

## Ethical considerations

The Senegalese National Ethical Committee of the Ministry of Health approved the surveillance protocol which lead to the obtention of human sera as less than minimal risk research, and written consent were not required. Throughout the study, the database was shared with the Epidemiology Department at the Senegalese Ministry of Health and Prevention for appropriate public health action.

## Reference

1. Mazzola LT, Kelly-Cirino C. Diagnostic tests for Crimean-Congo haemorrhagic fever: a widespread tickborne disease. BMJ Glob Health. 2019; 4(Suppl 2):e001114.

2. Ergonul O. Crimean-Congo hemorrhagic fever virus: new outbreaks, new discoveries. Curr Opin Virol. 2012; 2(2):215–220.

3. Calle-Prieto F de la, Martín-Quirós A, Trigo E, et al. Therapeutic management of Crimean-Congo haemorrhagic fever. Enferm Infecc Microbiol Clin (Engl Ed). 2018; 36(8):517–522.

4. Flusin O, Iseni F, Rodrigues R, et al. [Crimean-Congo hemorrhagic fever: basics for general practitioners]. Med Trop (Mars). 2010; 70(5–6):429–438.

5. Whitehouse CA. Crimean-Congo hemorrhagic fever. Antiviral Res. 2004; 64(3):145–160.

6. Papa A, Marklewitz M, Paraskevopoulou (Σοφία Παρασκευοπούλου) S, et al. History and classification of Aigai virus (formerly Crimean–Congo haemorrhagic fever virus genotype VI). Journal of General Virology. Microbiology Society,; 2022; 103(4):001734.

7. Camicas J-L, Gonzalez J-P, Leguenno B, et al. Recherches menées conjointement par l’USAMRIID, L’IPD et l’ORSTOM sur l’épidémiologie de la fièvre hémorragique de Crimée Congo au Sénégal et en Mauritanie. 1989; .

8. Tall A, Sall AA, Faye O, et al. Deux cas de fièvre hémorragique de Crimée-Congo (FHCC) contractée au Sénégal, en 2004, par des résidentes temporaires. Bull Soc Pathol Exot. 2009; :3.

9. Saluzzo JF, Aubry P, Aubert H, Digoutte JP. [Crimean-Congo hemorrhagic fever in Africa. Apropos of a case with hemorrhagic manifestations in Mauritania]. Bull Soc Pathol Exot Filiales. 1985; 78(2):164–169.

10. Weidmann M, Faye O, Faye O, et al. Development of mobile laboratory for viral haemorrhagic fever detection in Africa. 2018; :27.

11. Katoh K, Misawa K, Kuma K, Miyata T. MAFFT: a novel method for rapid multiple sequence alignment based on fast Fourier transform. Nucleic Acids Res. 2002; 30(14):3059–3066.

12. Nguyen L-T, Schmidt HA, Haeseler A von, Minh BQ. IQ-TREE: a fast and effective stochastic algorithm for estimating maximum-likelihood phylogenies. Mol Biol Evol. 2015; 32(1):268–274.

13. Sene O, Sagne SN, Ngom D, et al. Emergence of Crimean–Congo Hemorrhagic Fever Virus in Eastern Senegal in 2022. 2024; .

14. Sall AA, Zanotto PMDA, Sene OK, et al. Genetic Reassortment of Rift Valley Fever Virus in Nature. J Virol. 1999; 73(10):8196–8200.

15. Bowen MD, Trappier SG, Sanchez AJ, et al. A reassortant bunyavirus isolated from acute hemorrhagic fever cases in Kenya and Somalia. Virology. 2001; 291(2):185–190.

